# Patient-Reported Outcomes of Ugandans Living with Autoimmune Rheumatic Diseases

**DOI:** 10.1101/2020.12.30.20249043

**Authors:** Felix Bongomin, Maria Sekimpi, Barbra Natukunda, Anthony Makhoba, Mark Kaddumukasa

**Affiliations:** Department of Medicine, College of Health Sciences, Makerere University, Kampala, Uganda; Department of Immunology and Medical Microbiology, Gulu University Medical School, Gulu, Uganda; Department of Medicine, St. Francis’s Hospital- Nsambya, Kampala, Uganda; Department of Medicine, Mother Kevin Postgraduate Medical School, Uganda Martyrs University, Kampala, Uganda

**Keywords:** Autoimmune rheumatic diseases, SLE, rheumatoid arthritis, Health index, patient-reported outcomes, DMARDs, Ugand

## Abstract

**Purpose:** This study aimed to assess the patient–reported outcomes (PROs) in rheumatic patients attending two tertiary rheumatology clinics in Uganda.

**Methods:** A cross-sectional, clinical audit of patients aged 16 years or older with a confirmed diagnosis of rheumatic disease and receiving disease modifying anti-rheumatic drugs (DMARDs) was conducted between September and December 2020. Health index and overall self-rated health status were assessed using the ED-5D-5L tool. Comparisons for variables was performed using Student’s t-test or Mann-Whitney U for continuous numerical data while categorical data was compared using either *Χ*^2^ tests or Fisher’s exact tests as appropriate.

**Results:** We enrolled 74 eligible patients: 48 (64.9%) had rheumatoid arthritis (RA), 14 (18.9%) had systemic lupus erythematosus (SLE), and 12 (16.2%) had other autoimmune rheumatic disorders. Majority (n=69, 93.2%) were female with a mean ±SD age of 45 ± 17 years. Fourteen (18.9%) patients were on concomitant herbal medication while using DMARDs and 26 (35.1%) self-reported at least 1 adverse drug reactions to the DMARDS. Any level of problem was reported by 54 (72.5%) participants for mobility, 47 (63.5%) for self-care, 56 (75.6%) for usual activity, 66 (89.1%) for pain and discomfort, and 56 (75.6%) for anxiety/depression. Patients with SLE had higher median health index compared to those other autoimmune rheumatic disorders (p<0.0001). Overall self-rated health status was comparable across clinical diagnoses (p=0.2), but better for patients who received care from private (Nsambya Hospital) compared to public hospital (Mulago Hospital) (65 vs. 50, p=0.009).

**Conclusion:** There is a substantial negative impact of autoimmune rheumatic diseases on quality of life of patients, especially those receiving care from a public facility in Uganda.

**Clinical Significance:**

1. Adverse drug reactions to DMARDs was reported by more than one-third of the patients
2. SLE patients have better quality of life compared to patients with other autoimmune rheumatic disease.
3. Concomitant use of herbal medication is common and associated with lower health index and lower overall self-rated health status.
4. Autoimmune rheumatic diseases impose a heavy financial burden on affected patients, over 70% of the study patients required financial support for management of their disease and a high proportion of these patients were not on their DMARD therapy the week prior to their scheduled clinic appoints.

## Introduction

Autoimmune rheumatic disorders such as rheumatoid arthritis (RA) and systemic lupus erythematosus (SLE) are associated with pain, disability and several co-morbid conditions thus significantly impacting on quality of life and overall wellbeing of the affected individuals and their families.^1^ Consequently, higher morbidity and mortality rates are observed among these individuals compared to the general population^1,2^. Also, there’s a significant individual differences in the day-to-day variability of pain, fatigue, and well-being in patients with rheumatic disease.^3^ Therefore, quality of life is central in the care of patients with autoimmune rheumatic diseases and is an important target in therapeutic advances in rheumatology while evaluating or managing these patients with disease modifying anti-rheumatic drugs (DMARDs).^4^

Patient-reported outcomes (PROs) are patient’s perspectives on their disease activity, functional status, and quality of life^5^. Patient-reported outcome measures (PROMs), are a set of widely available tools directly capture PROs and are increasingly being used in clinical rheumatology practice and in research to help inform patient-centered care and clinical decision-making even among vulnerable rheumatic patients such as those with low health literacy or English proficiency.^6^

There are no locally validated rheumatic disease specific PROMs in Africa and data on PROs of patients with rheumatic diseases in Africa is scanty, even though these diseases, especially RA and SLE are increasingly being reported in Africa.^7–9^ This study aimed to describe PROs of patients with autoimmune rheumatic diseases in two tertiary care centers in Uganda.

## Patients and Methods

### Study design and settings

This descriptive, cross-sectional clinical audit recruited consecutive outpatients attending two rheumatology clinics at Mulago National Referral Hospital (Mulago Hospital), Kampala, Uganda and St. Francis’s Hospital-Nsambya, Kampala, Uganda (Nsambya Hospital) between September and December 2020. Mulago Hospital, located in the capital city, Kampala, is the largest public health facility in Uganda serving as a national super-specialized hospital with over 1,000-bed capacity. Nsambya Hospital is a faith-based, private-not-for-profit hospital also located in Kampala, Uganda.

### Study population

Patients aged 16 years or older with a diagnosis of an autoimmune rheumatologic disease diagnosed by one of the two experienced rheumatologists (AM and MK) for whom at least one of the DMARDs was prescribed in their last previous clinic visit constituted the study population.

### Data collection

Data were collected using semi-structured questionnaires administered by the treating physicians (the authors) during routine clinical care. This audit was anonymous, consisted of semi-structured questions, which were available only in English. Data was collected on the following parameters: (1) patient socio-demographic characteristics: age, gender, marital status, level of education, current employment status, monthly income and financial support from family members; (2) Clinical diagnosis: duration of illness, self-reported disease severity, disease flares, hospitalization and family history of autoimmune disease; (3) Medication: DMARDs used, duration of therapy, source of DMARDs, monthly expenditure on DMARDs, satisfaction with treatment, concomitant use of herbal medication, adverse drug reactions; (4) Number of additional medications used daily; and (5) co-morbidities

### Patient-reported outcome measure

The EQ-5D-5L, a standardized instrument for use as a measure of health outcomes consisting of 5 dimensions and 5 levels was administered to the participants^10^. The tool has been previously used in sub-Saharan Africa and is being validated in Ethiopia^11,12^. The 5 dimensions assessed were mobility, self-care, usual activities, pain/discomfort, and anxiety/depression. Each dimension has five levels (no problems, slight problems, moderate problems, severe problems, extreme problems/unable to). Health state profile was generated from these dimensions and levels. Overall self-rated health status was assessed using the visual analogue scale (VAS) on which the patient rates his/her perceived health from 0 (the worst imaginable health) to 100 (the best imaginable health).

### Data analysis

Baseline characteristics were summarized using medians and ranges or means and standard deviations (SD) for continuous variables and frequencies and percentages for categorical variables. Comparisons for variables were performed using Student’s t-test or Mann-Whitney U (for two group comparisons) and the one-way analysis of variance or Kruskall-Wallis (for more than two group comparisons) for continuous numerical data. Categorical data were compared using either *Χ*^2^ tests or Fisher’s exact tests as appropriate. Health state index scores generally range from less than 0 (where 0 is the value of a health state equivalent to dead; negative values representing values as worse than dead) to 1 (the value of full health) were calculated from individual health profiles using crosswalk value sets for Zimbabwe (Reference). Statistical analyses were performed using STATA 16.0 and GraphPad Prism 8.0. A p<0.05 was considered to indicate statistical significance.

### Ethical considerations

This was a clinical audit, evaluating standard of care of patients in our clinics. All patients provided written informed consent and the study protocol was in compliance with the ethical guidelines of the *Declarations of Helsinki*.

## Results

### Sociodemographic characteristics

A total of 74 eligible patients were studied: 41 (55.4%) from Mulago Hospital and 33 (44.6%) from Nsambya Hospital. Majority were female (n=69, 93.2%) with a mean ±SD age of 45 ± 17 years. Thirty-one (41.9%) patients were single, 29 (39.1%) had received tertiary education and 49 (66. 2%) were not formally employed. The median (range) monthly income was 300,000 (30,000 – 1,000, 000) Ugandan shillings (UGX). Fifty-five (74.3%) participants received financial support from their children (n=18, 24.3%), other family members (n=15, 20.3%), their spouses (n=12, 16.2%), their parents (n=7, 9.5%) or from the church (n=3, 4.1%), **Table 1**.

**Table 1:**
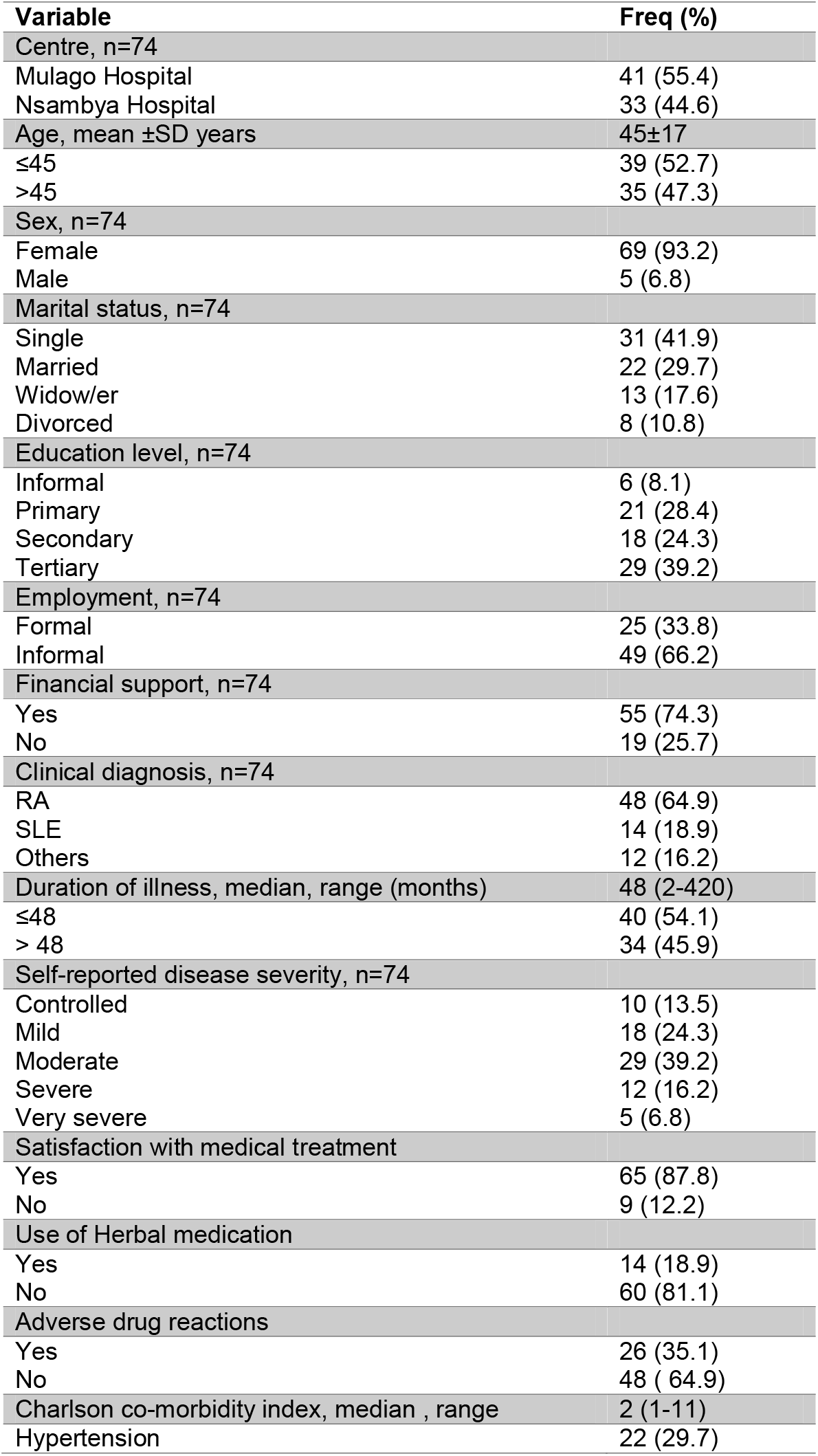
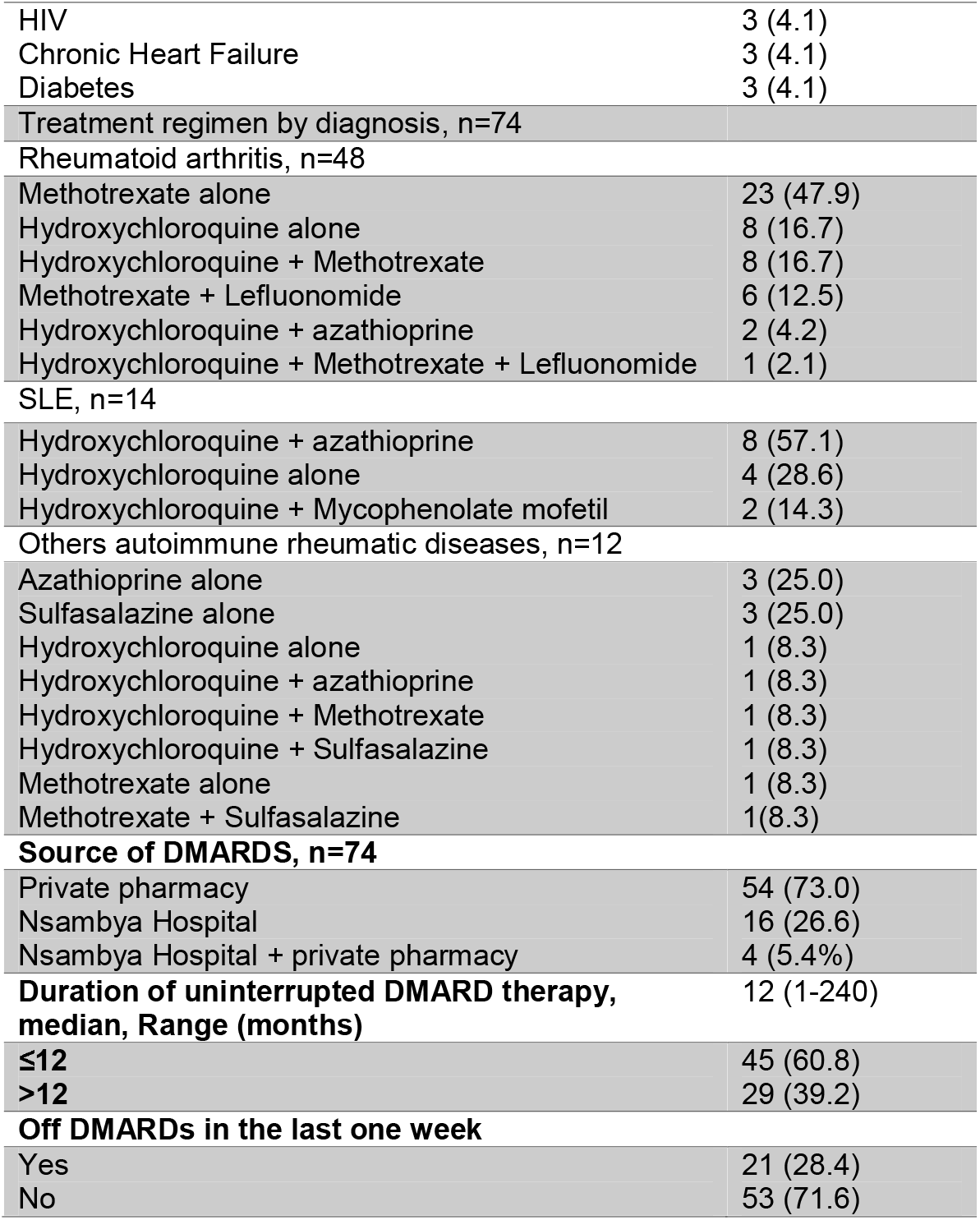
Sociodemographic and clinical characteristics of the participants.

### Clinical characteristics

Forty-eight (64.9%) patients had RA, 14 (18.9%) had SLE, and 12 (16.2%) had other rheumatic disorders namely spondyloarthropathy (n=5), systemic sclerosis (n=3), juvenile idiopathic arthritis (n=2), and inflammatory myopathy (n=2). The median (range) duration of illness was 48 (2-420) months. On the day of the clinic visit, patients’ self-rated severity of their illnesses was as follows: Controlled (n=10, 13.5%), mild (n=18, 24.3%), moderate (n=29, 39.2%), severe (n=12, 16.2%), and very severe (n=5, 6.8%). The median (range) episodes of disease flares in the past 3 months was 1 (range: 0-20). Thirty-two (43.2%) patients had at least one co-morbidity. Of these, 23 (71.9%) were RA patients, 4 (12.5%) were SLE and 5 (15.6%) were patients who had other clinical diagnoses.

### Disease modifying anti-rheumatic drug therapy

None of the patients was on biologic DMARDs. Majority of the patients with RA were on monotherapy of methotrexate (n=23, 47.9%), those with SLE were mostly either on monotherapy of hydroxychloroquine or in a combination with azathioprine (n= 12, 85.7%), and half of patients with other rheumatic diseases were either on azathioprine or sulfasalazine (n=6, 50%), **Table 1**. The median duration of DMARD therapy was 12 (range: 1-24) months. All 41 patients from Mulago Hospital bought their DMARDs from private pharmacies. On the other hand, 16/33 patients from Nsambya Hospital bought their DMARDS from the hospital pharmacy, 13/33 from private pharmacies and 4 from either private pharmacies or Nsambya Hospital pharmacy. The median (range) monthly cost of DMARDs was 120,000 (12,800 – 2,000,000) UGX. Sixty-four (86.4%) participants reported satisfaction with DMARD treatment. However, 14 (18.9%) participants were on concomitant herbal medication while using DMARDs. Twenty-six (35.1%) participants reported at least 1 adverse drug reactions (ADRs) to the DMARDs. Most ADRs were observed with methotrexate (10/26; 4 patients reported dizziness, 3 weakness, 2 gastrointestinal (GI) disturbances and 1 pulmonary fibrosis), hydroxychloroquine (8/26: 1 visual impairment, 2 rashes, and 5 dizziness), sulfasalazine (4/26: 1 nightmare, 3 GI disturbance), azathioprine (3/26; all 3 reported weakness), and 1 patient reported diarrhea while on mycophenolate mofetil.

Twenty-one (28.4%) patients, most of whom were attending Mulago Hospital Rheumatology Clinic (18/21 (86%) vs. 3/21 (14%), p=0.01) were off DMARDs in the week prior to clinic visit.

### Health profiles and overall health status

Table 3 and Table 4 summarizes the health indices and overall self-rated health status of the participants across sociodemographic and clinical characteristics. Regarding the health profiles of the participants, 71 (96%) participants reported at least one activity limitation. Any level of problem was reported by 54 (72.5%) participants for mobility, 47 (63.5%) for self-care, 56 (75.6%) for usual activity, 66 (89.1%) for pain and discomfort, and 56 (75.6%) for anxiety/depression. The median health index was 0.675 (range: 0.163 – 0.9). Both health index and overall self-rated health status were higher for patients who received care from Nsambya Hospital compared to Mulago Hospital (0.718 vs. 0.585, p<0.0001) and (63.6 vs. 53.7, p<0.0001), respectively **Table 3 and 4**. Also, patients with SLE had higher median health index compared to those with RA or other rheumatic disease, **Table 3**. Patients who reported controlled or mild disease, and those who reported satisfaction with medical (DMARDs) therapy had higher health indices and higher overall self-rated health status (all p-values <0.001), **Table 3 and 4**. Conversely, patients who reported concomitant herbal medication use had lower health index (0.558 vs. 0.664, p=0.03) and lower overall self-rated health status (47.6 vs. 60.5, p=0.01) compared to those who did not report use of herbal medications, **Table 3 and 4**. There was a trend towards patients with co-morbidities having a better overall self-reported health status compared to those without co-morbidities (58.6 vs. 57.7, p=0.05), **Table 4**.

**Table 2:** EQ-5D-5L frequencies and proportions reported by dimension and level.

|  | MOBILITY<br>N (%) | SELF-CARE<br>N (%) | USUAL ACTIVITIES<br>N (%) | PAIN/DISCOMFORT<br>N (%) | ANXIETY/DEPRESSION<br>N (%) |
| --- | --- | --- | --- | --- | --- |
| LEVEL 1<br>NO PROBLEMS | 20 (27.0) | 27 (36.5) | 18 (24.3) | 8 (10.8) | 18 (24.3) |
| LEVEL 2<br>SLIGHT PROBLEMS | 26 (35.1) | 23 (31.1) | 26 (35.1) | 23 (31.1) | 26 (35.1) |
| LEVEL 3<br>MODERATE PROBLEMS | 15 (20.3) | 15 (20.3) | 20 (27.0) | 26 (35.1) | 20 (27.0) |
| LEVEL 4<br>SEVERE PROBLEMS | 12 (16.2) | 8 (10.8) | 6 (8.1) | 17 (23) | 8 (10.8) |
| LEVEL 5<br>EXTREME<br>PROBLEMS/UNABLE TO DO | 1 (1.4) | 1 (1.4) | 4 (5.4) | 0 (0) | 2 (2.7 ) |
| TOTAL | **74 (100)** | **74 (100)** | **74 (100)** | **74 (100)** | **74 (100)** |

**Table 3:** Health indices of the participants across sociodemographic and clinical characteristics.

| Variable | Freq (%) | Health index<br>Mean (SD) | p-value |
| --- | --- | --- | --- |
| **ALL, n=74** | 74 (100%) | 0.644 ± 0.164 | - |
| Centre, n=74 |  |  |  |
| Mulago Hospital | 41 (55.4) | 0.585 ± 0.177 | <0.0001 |
| Nsambya Hospital | 33 (44.6) | 0.718 ± 0.110 |  |
| Age, year |  |  |  |
| ≤45 | 39 (52.7) | 0.657 ± 0.138 | 0.516 |
| >45 | 35 (47.3) | 0.632 ± 0.185 |  |
| Sex, n=74 |  |  |  |
| Female | 69 (93.2) | 0.637 ± 0.162 | 0.160 |
| Male | 5 (6.8) | 0.744 ± 0.177 |  |
| Marital status, n=74 |  |  |  |
| Single | 31 (41.9) | 0.673 ± 0.169 | 0.633 |
| Married | 22 (29.7) | 0.629 ± 0.144 |  |
| Widow/er | 13 (17.6) | 0.611 ± 0.212 |  |
| Divorced | 8 (10.8) | 0.628 ± 0.105 |  |
| Education level, n=74 |  |  |  |
| Informal | 6 (8.1) | 0.675 ± 0.139 | 0.227 |
| Primary | 21 (28.4) | 0.594 ± 0.162 |  |
| Secondary | 18 (24.3) | 0.625 ± 0.169 |  |
| Tertiary | 29 (39.2) | 0.686 ± 0.162 |  |
| Employment, n=74 |  |  |  |
| Formal | 25 (33.8) | 0.672 ± 0.172 | 0.302 |
| Informal | 49 (66.2) | 0.630 ± 0.159 |  |
| Financial support, n=74 |  |  |  |
| Yes | 55 (74.3) | 0.637 ± 0.161 | 0.528 |
| No | 19 (25.7) | 0.665 ± 0.665 |  |
| Clinical diagnosis, n=74 |  |  |  |
| RA | 48 (64.9) | 0.601 ± 0.173 | 0.007 |
| SLE | 14 (18.9) | 0.738 ± 0.121 |  |
| Others | 12 (16.2) | 0.704 ± 0.097 |  |
| Duration of illness, median, range (months),<br>, n=74 | 48 (2-420) | 0.644 ± 0.164 |  |
| ≤48 | 40 (54.1) | 0.682 ± 0.125 | 0.029 |
| > 48 | 34 (45.9) | 0.599 ± 0.193 |  |
| Self-reported disease severity, n=74 |  |  |  |
| Controlled | 10 (13.5) | 0.740 ± 0.112 | <0.0001 |
| Mild | 18 (24.3) | 0.719 ± 0.120 |  |
| Moderate | 29 (39.2) | 0.650 ± 0.158 |  |
| Severe | 12 (16.2) | 0.467 ± 0.090 |  |
| Very severe | 5 (6.8) | 0.572 ± 0.234 |  |
| Satisfaction with medical treatment |  |  |  |
| Yes | 65 (87.8) | 0.668 ± 0.145 | 0.001 |
| No | 9 (12.2) | 0.472 ± 0.194 |  |
| Use of Herbal medication |  |  |  |
| Yes | 14 (18.9) | 0.558 ± 0.231 | 0.03 |
| No | 60 (81.1) | 0.664 ± 0.139 |  |
| Adverse drug reactions |  |  |  |
| Yes | 26 (35.1) | 0.627 ± 0.181 | 0.516 |
| No | 48 ( 64.9) | 0.653 ± 0.155 |  |
| Co-morbidity |  |  |  |
| Yes | 32 (43.2) | 0.642 ± 0.170 | 0.91 |
| No | 42 (56.8) | 0.646 ± 0.161 |  |
| Source of DMARDS, n=74 |  |  |  |
| Private pharmacy | 54 (73.0) | 0.644 ± 0.164 | 0.196 |
| Nsambya Hospital | 16 (26.6) | 0.709 ± 0.099 |  |
| Nsambya Hospital + private pharmacy | 4 (5.4%) | 0.649 ± 0.102 |  |
| **Duration of uninterrupted DMARD therapy, median, Range (months), n=74** | 12 (1-240) | 0.646 ± 0.161 |  |
| **≤12** | 45 (60.8) | 0.675 ± 0.133 | 0.042 |
| **>12** | 29 (39.2) | 0.594 ± 0.195 |  |
| **Off DMARDs in the last one week** |  |  |  |
| Yes | 21 (28.4) | 0.622 ± 0.185 | 0.771 |
| No | 53 (71.6) | 0.652 ± 0.180 |  |

**Table 4:**
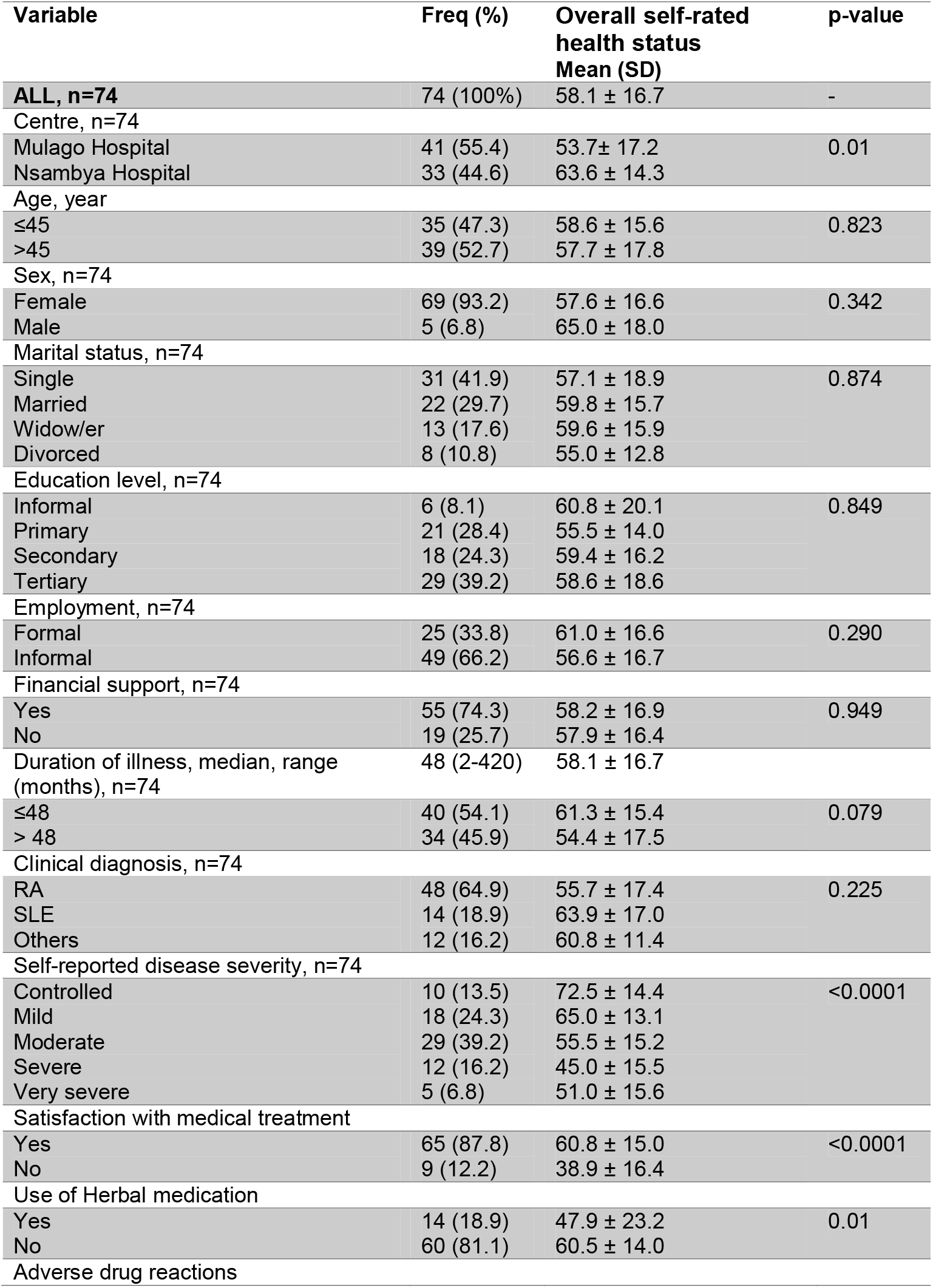
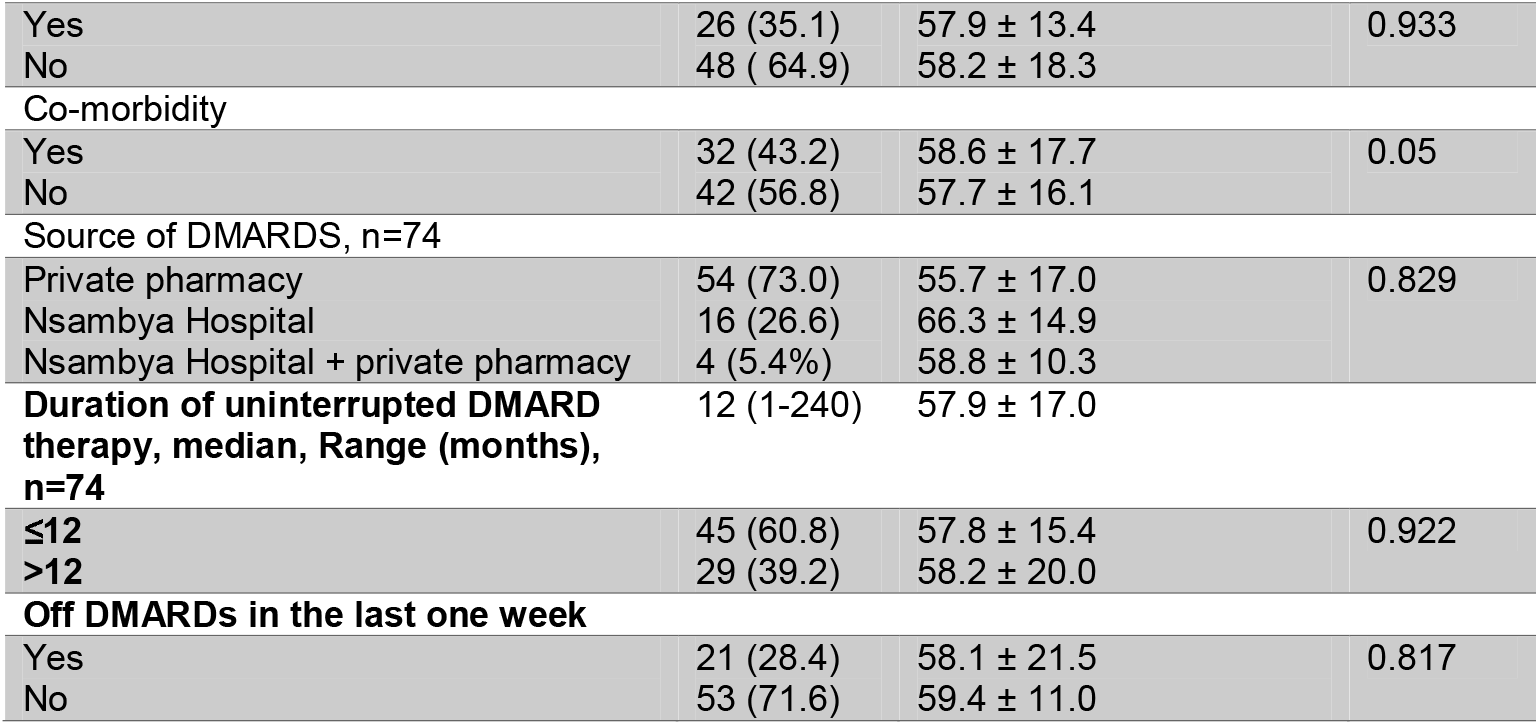
Overall self-rated health status of the participants across sociodemographic and clinical characteristics.

## Discussion

Understanding PROs influence treatment decisions and inform clinical care in patients with autoimmune rheumatic disease.^13,14^ In the present study, among Ugandan patients with autoimmune rheumatic diseases, over 95% of the patients reported at least one activity limitation. This finding is consistent with the 2020 American College of Rheumatology (ACR) patients survey, where about 83% of people living with a rheumatic disease reported at least one activity limitation as a result of their disease, including ability to exercise, work, and perform physical activities.^15^ Our findings suggest that patients with SLE have a better quality of life compared to patients with other autoimmune rheumatic diseases which is in line with prior investigation.^16^ Contrastingly, a recent study from Kenya showed that patients with SLE had significantly low health-related quality of life.^17^ This is probably because the Kenyan patients had more severe disease, though much younger age than our population. Remarkably, participants with duration of illness of 4 years or less and those who were on DMARDs for less than 1 year had higher health indices. Equally remarkably, overall self-rated health status was comparable across groups and sub groups of illness duration and duration of uninterrupted DMARDs therapy.

Age, disease severity and co-morbidities are important predictors of quality of life of patients with autoimmune diseases.^17–19^ Thus it was not surprising that in our study, patients with controlled or mild disease and those who reported satisfaction with DMARDS had higher health indices and high self-rated health status. Current rheumatic management guidance emphasizes the treat-to-target approach, as patients in remission or low disease activity tends to have a better quality of life indices^14^. However, access and affordability of both conventional and biologic DMARDs remains a challenge worldwide.^15,20^ indeed, none of our study participants was on a biologic DMARDS. Lack of access to and non-affordability of DMARDs have negative association with disease activity and a poorer quality of life.^20^ This is evident in our study where patients attending care in a private hospital with better access to DMARDs had better health indices and overall self-rated health status.

DMARDs are expensive and are unaffordable by most patients. In the 2020 ACR patient survey, the median annual out-of-pocket spending on treatment for rheumatic disease was $1,000 per year.^15^ On average, out-of-pocket expenditure on DMARDs of our patients was about $400 per year. This is quite high and explains the high proportion of patients not being on their DMARDs the week prior to their scheduled clinic appoints. In Uganda, much as DMARDs such as methotrexate are on the essential medicine list, they are not routinely available for the care of patients with rheumatic diseases. The heavy financial burden of these diseases and their management explains the huge need for financial support observed in over 70% of our patients. Consequently, patients who are wealthier or have health insurance services are able to access these medicines in private settings and have better adherence and health outcomes as observed in one of the centres in the present study.

One in every 5 patients with rheumatic disease in our cohort reported concomitant use of herbal medication. Regrettably, this was associated with lower health index and lower overall self-rated health status. Despite the fact that ADRs were similar among those who were on herbal medications and those not using herbs, these findings should encourage clinicians to always assess for herbal medication use among these patients and provide appropriate counseling. However, it is unclear whether the poor quality of life of patients on concomitant herbal medication was truly due to negative impacts of herbal medicines on rheumatic diseases or because patients who showed poor response while on DMARDs had uncontrolled disease and therefore sought for herbal remedy for a better disease control. Herbal medication use remains an area of further research among these patients. Known beneficial add-on therapy in patients with rheumatic diseases revolves around optimization of the management of underlying co-morbidities, physical and occupational therapies.^15^

Our study has some important limitations. Firstly, we were unable to assess disease specific severity for the different rheumatic diseases. However, we were able to elicit patients-reported disease severity which fairly correlates with disease severity scores. Secondly, we were unable to formally assess for medication adherence using validated tools due to lack of access to license. Thirdly, we were unable to use disease specific health-related outcome measures such as LupusQoL.^16^ However, ED-5D-5L has been shown to be a reliable tool for these group of patients.^21^ Lastly,. measurements of test–retest reliability were not done because patients were assessed on only one clinic visit. However, this the first study from Uganda and one of the few in the region to report on quality of life of patients with autoimmune rheumatic diseases receiving DMARDs. Future studies would aim at correlating health indices with disease severity and medication adherence in our setting. At policy level, we need to identify strategies to widely increase availability; accessibility and affordability of DMARDs in Uganda should be explored. It’s timely to welcome clinical trials on biologic DMARDs for our patients to evaluate short- and long-term outcomes.

## CONCLUSION

In conclusion, over 95% of Ugandan patients with autoimmune rheumatic diseases on DMARDs have at least one activity limitation. SLE patients have better quality of life compared to patients with RA or rheumatic disease. Concomitant use of herbal medication is common and associated with lower health index and lower overall self-rated health status.

## Data Availability

All relevant data have been included in the manuscript

## Acknowledgement

Mark Kaddumukasa is supported by a grant from the National Institutes Health (K43TW010401 NINDS and Fogarty International Center (FIC). We are indebted to our patients who have become a significant part of us and through whom we are able to enjoy the practice of clinical rheumatology. We appreciate the administrative support from the study sites.

## Authors’ contribution

All authors significantly contributed to the conceptualization, design, data collection, analysis and interpretation and drafting of the manuscript. The final manuscript was read and approved by all authors.

